# Design and Rationale of the APOLLO Registry: Integrated Assessment of Cardio-Renal Perfusion and Cardiovascular-Kidney-Metabolic Phenotyping Using ^82^Rb-PET/CT

**DOI:** 10.64898/2026.10.01.26364477

**Authors:** Begawan A. Siadari, Melina C. den Haan, Frits Smit, Sandra de Boer, Kjell Bogaard, Etto C. Eringa, Thomas Ruytenberg, Floris H.P. van Velden, Lioe-Fee de Geus-Oei, Ilona A. Dekkers

## Abstract

**Background:** Cardiovascular-kidney-metabolic (CKM) syndrome encompasses interconnected diseases that share pathophysiological mechanisms, including microvascular dysfunction. Although rubidium-82 positron emission tomography/computed tomography (²Rb-PET/CT) is routinely used for myocardial perfusion imaging, its potential for integrated cardio-renal perfusion assessment and broader CKM phenotyping remains underexplored. We recently developed a compartment modelling approach to quantify renal perfusion from existing routine cardiac ^82^Rb-PET/CT scans, enabling simultaneous assessment of myocardial and kidney perfusion. However, real-world registries integrating these phenotypes with clinical outcomes are lacking.

**Aim:** To establish a registry integrating ^82^Rb-PET-derived myocardial and renal perfusion, CT-derived CKM imaging biomarkers, and detailed clinical and outcome data.

**Methods:** The APOLLO registry is a single-centre, retrospective database comprising approximately 1,200 consecutive patients who underwent myocardial perfusion imaging with ^82^Rb-PET/CT at Alrijne Hospital (Leiderdorp, the Netherlands) between 2021-2023. Clinical, laboratory, medication, echocardiographic, and imaging data were retrospectively collected. Myocardial perfusion was quantified using myocardial blood flow and myocardial flow reserve, and renal perfusion using renal blood flow and effective renal plasma flow. Additional CT-derived biomarkers included measures of adiposity and frailty, great vessel diameters, and vascular calcification. Echocardiographic diastolic dysfunction grade, H_2_FPEF score, CKM stage, and major adverse cardiovascular events (MACE) were also collected.

**Conclusion:** The APOLLO registry integrates ²Rb-PET-derived myocardial and renal perfusion measurements with CT-derived CKM imaging biomarkers and clinical outcomes. This enables investigation of cardio-renal perfusion and their associations with CKM phenotypes and cardiovascular outcomes.

## Introduction

Cardiovascular, kidney, and metabolic diseases frequently coexist and collectively account for a substantial proportion of global morbidity and mortality [1]. These conditions have been studied as separate disease entities; however, growing evidence demonstrated a shared common risk factors and pathophysiological pathways reflected in the Cardiovascular-kidney-metabolic (CKM) framework, proposed by the American Heart Association (AHA) [2,3]. Microvascular dysfunction has been proposed as an important component in this interconnected disease spectrum [3]. Coronary microvascular dysfunction (CMD) is increasingly recognized as an important contributor to heart failure with preserved ejection fraction (HFpEF), obesity, diabetes, and chronic kidney disease (CKD), which has been linked with adverse cardiovascular outcomes [4]. Similarly, alterations in the renal microcirculation may contribute to kidney dysfunction and cardio-renal disease progression [5,6]. Despite the growing recognition of microvascular dysfunction importance across CKM spectrum, integrated in vivo assessment of cardiac and renal microvascular function remains limited.

Positron emission tomography/computed tomography using the perfusion tracer rubidium-82 (^82^Rb-PET/CT) is an established method for myocardial perfusion imaging. Quantitative assessment includes with myocardial blood flow (MBF) at rest and during pharmacological stress, as well as myocardial flow reserve (MFR), which are key parameters for detection of myocardial ischemia [7–9]. However, ²Rb-PET/CT may also provide quantitative imaging biomarkers for integrated cardio-renal perfusion assessment and broader CKM phenotyping. In this context, we recently explored the potential of repurposing routinely acquired cardiac ²Rb-PET/CT scans to retrospectively quantify renal perfusion. This led to the development of a kidney-specific compartmental modelling approach that enables quantification of myocardial and renal perfusion from a single cardiac ²Rb-PET/CT examination, without the need for additional imaging [10]. Furthermore, the accompanying low-dose CT scan, acquired for PET attenuation correction, can be used to derive additional imaging biomarkers relevant to the CKM framework, including measures of adiposity, vascular calcification, great vessel diameters, and frailty [11].

Despite these opportunities, real-world registries that integrate PET-derived myocardial and renal perfusion, CT-derived CKM imaging biomarkers, and clinical data are lacking. Such registries with comprehensive phenotyping across the CKM spectrum may facilitate investigation of the relationship between cardio-renal perfusion, cardiometabolic risk, and clinical outcome. To address this gap, we established the APOLLO Registry, a large-scale retrospective cohort comprising approximately 1,200 patients who underwent cardiac ²Rb-PET/CT. The design and rationale of the registry are described in this paper.

## Methods

### Registry design

The APOLLO registry is a single-centre retrospective observational database of patients who underwent myocardial perfusion imaging of suspected myocardial ischemia at the Alrijne Hospital (Leiderdorp, the Netherlands). Data collection commenced in January 2025 and is still ongoing. Clinical, imaging, and outcome data are collected and managed within Castor Electronic Data Capture (EDC) [12] according to FAIR (Findable, Accessible, Interoperable, and Reusable) data principles. Ethical approval for this study was obtained from the Medical Research Ethics Committee Leiden Den Haag Delft (METC LDD; nWMODIV2_20240025). This registry was deemed not subjected to the Dutch Medical Research Involving Human Subjects Act (Wet Medisch-wetenschappelijk Onderzoek met mensen, WMO), as participants were not exposed to additional study-related procedures or questionnaires. The requirement for written informed consent was waived due to the retrospective nature of the study, the use of pseudonymized data, and the large study population of 1,174 patients.

### Study population

Consecutive patients who underwent cardiac ^82^Rb-PET/CT between January 2021 and December 2023 at Alrijne hospital constituted the APOLLO registry cohort. Eligible patients had to have sufficient kidney coverage within the scan range of the ^82^Rb-PET (>15 consecutive slices or >5.7cm coverage of the upper pole of at least one kidney). In addition, patients need to have clinical and laboratory data available within 1 year of the ^82^Rb-PET/CT examination, and a rejected-to-accepted beat ratio below 0.25. Data collection of the included 1,174 patients in the larger APOLLO registry is currently ongoing.

### Imaging acquisition

Dynamic rest and stress ^82^Rb-PET/CT examinations were performed on a Discovery MI 5-ring PET/CT system (GE HealthCare). The imaging protocol consisted of a low-dose CT acquisition used for attenuation correction, a dynamic ECG-gated PET acquisition, and, in a subset of patients, an additional ECG-gated CT for coronary artery calcium scoring (figure 1). All PET acquisitions were performed at rest and during pharmacological stress, induced using adenosine (140 µg/kg/min over 6 min) or regadenoson (400 μg, independent of patient’s weight). More details on the acquisition and subsequent image reconstruction are provided in the Supplemental Methods (Figure S1).

**Figure 1.**
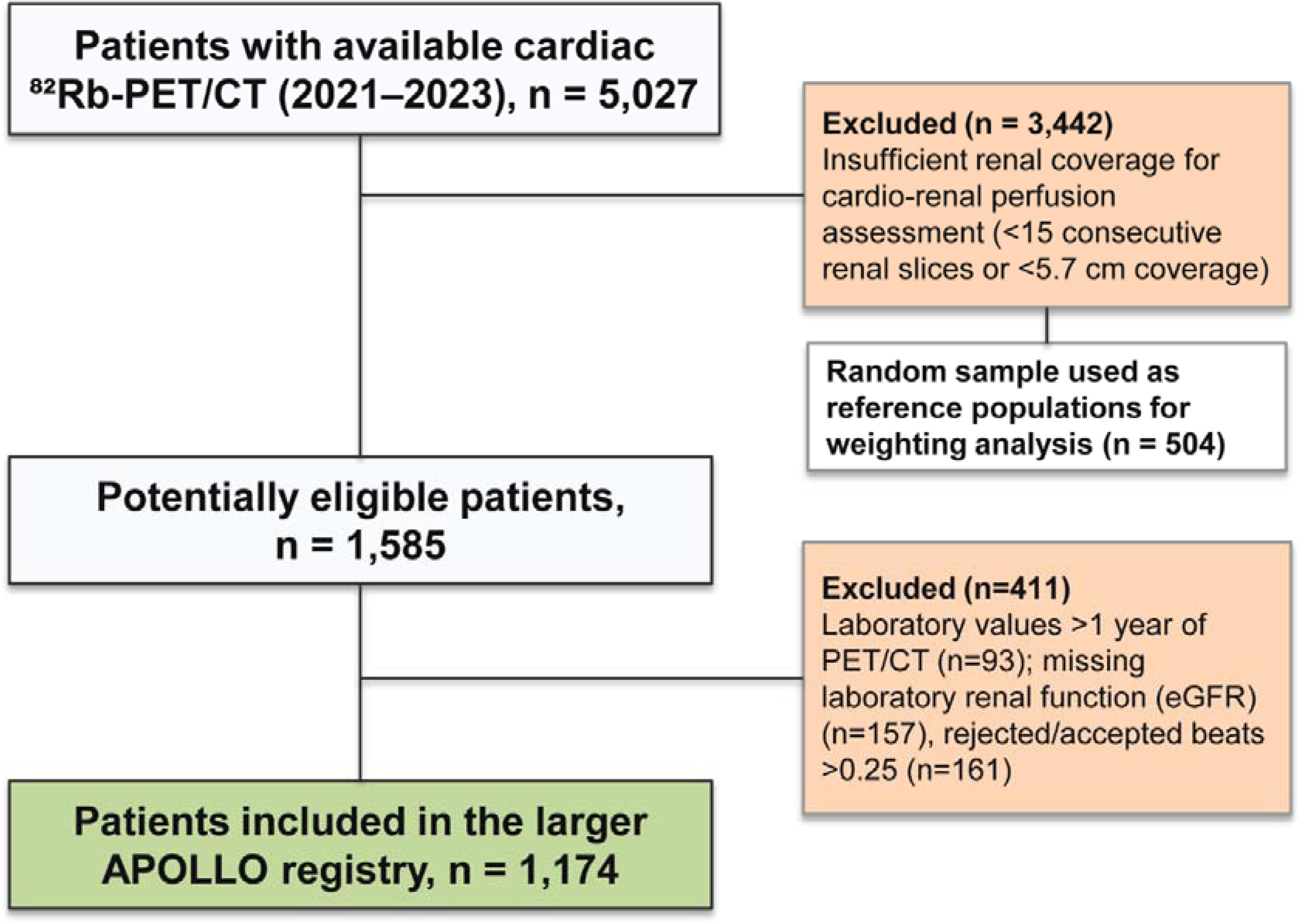
Flow chart of the APOLLO registry. CT: Computed Tomography; PET: Positron Emission Tomography; 82Rb: Rubidium-82; eGFR: estimated glomerular filtration rate

### Collection of PET-derived hemodynamic measures

Dynamic PET datasets were analysed semi-automatically for cardiac [9] and renal [10] perfusion using Corridor4DM software (INVIA Medical Imaging Solution) and PMOD software (PMOD Technologies Ltd) respectively. Detailed descriptions of image processing, region of interest (ROI) placement, kinetic modelling, and perfusion quantification are provided in the Supplemental Methods.

#### Cardiac perfusion

Primary cardiac perfusions metrics collected were MBF at rest and during pharmacological stress (ml/min/g) and MFR, defined as the ratio of stress MBF to rest MBF [8,9]. Integrated myocardial flow reserve (iMFR) maps were generated by combining relative perfusion and MFR to characterize focal and diffuse coronary microvascular dysfunction for each coronary artery territory and in total (Figure 2B). Global MFR <2.0 was used as marker of impaired vasodilator capacity. Reduced global MFR may also reflect obstructive epicardial disease, CMD was defined using the iMFR classification as diffusely impaired flow (MFR < 2.0 without relative perfusion defect). Patients with obstructive CAD were retained and distinguished by iMFR rather than excluded [9].

**Figure 2.**
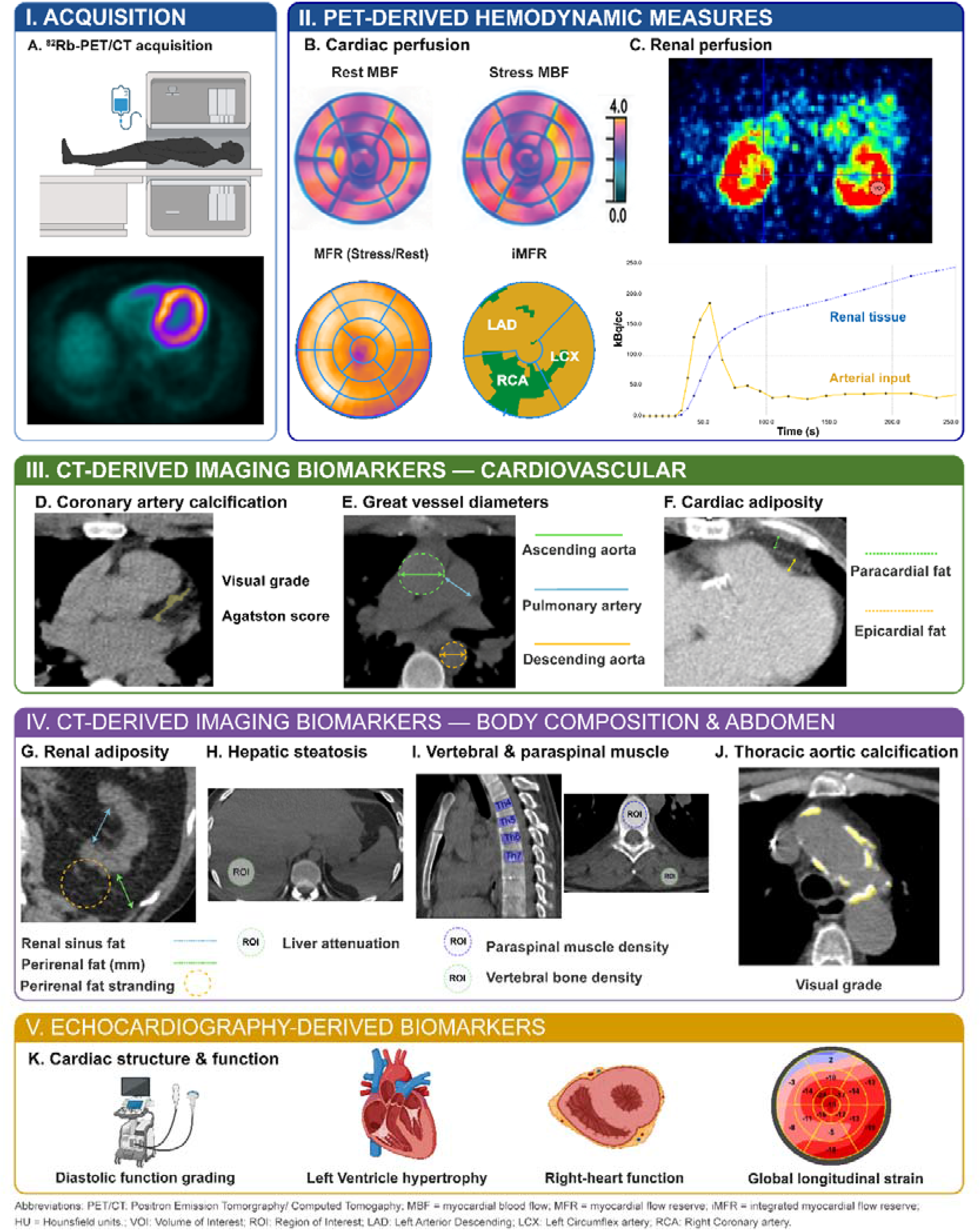
Overview of Imaging biomarker retrospectively collected for the APOLLO registry. (A) Acquisition and representation (axial view) of dynamic ^82^Rb-PET/CT scan. (B) Cardiac perfusion (ml/min/g) derived from PET/CT for MBF at rest and stress, MFR, and iMFR (in LAD, RCA, and LCX) using Corridor4DM software. (C) Renal perfusion (ml/min/100g) derived from PET/CT to create time-activity curve using PMOD software. (D) Visual grading of coronary artery calcification (Absent-limited, moderate, severe) and Agatston score collection. (E) Great Vessel diameters (mm) for pulmonary artery (blue), ascending (green), and descending aorta (yellow). (F) Measurement of cardiac adiposity width (mm) in epicardial (yellow) and paracardial (green) taken at LAD level. (G) Measurement of perirenal (green) fat width (mm), visual grading of renal sinus (blue) fat (absent-limited, low, moderate, high) and perirenal (yellow) fat stranding (low, intermediate, high). (H) Hepatic steatosis (HU) measurement with ROI in liver segment VI and spleen. (I) Vertebral and paraspinal muscle density (HU) measurement taken at TH4 to TH7 level, with ROIs located in vertebral body and paraspinal muscles. (J) Visual grading of Thoracic aorta calcification (Absent-limited, mild, moderate, severe). (K) Echocardiographic-derived measurements for cardiac structure and function utilized for H_2_FPEF score. PET/CT: Positron Emission Tomorgraphy/ Computed Tomogaphy; MBF = myocardial blood flow; MFR = myocardial flow reserve; iMFR = integrated myocardial flow reserve; Ca: Vascular Component; Ck: Tissue/ Renal Compartment HU = Hounsfield units.; VOI: Volume of Interest; ROI: Region of Interest; LAD: Left Arterior Descending; LCX: Left Circumflex artery; RCA: Right Coronary artery.

#### Renal perfusion

Renal perfusion was derived from the available upper pole of the kidney within the scan range of the dynamic PET acquisition at rest [10]. Renal blood flow (RBF; ml/min/100g) was quantified from influx rate constant (K1) values using a one-compartment kinetic model (Figure 2C) [10,15,16]. Effective renal plasma flow (ERPF, mL/min) was calculated using available haematocrit (Ht) values; using the formula ERPF= RBF * (1-Ht).

### Collection of CT-derived CKM imaging biomarkers

CT-derived CKM imaging biomarkers were categorized into biomarkers of adiposity, great vessel diameter, vascular calcification, frailty, and renal inflammation. Measurements of CT-derived CKM imaging biomarkers were performed by a single observer (B.S.) according to predefined protocols and reviewed by an experienced radiologist (I.D.), and robustness was evaluated in a predefined subset of scans (Supplemental methods).

#### Adiposity biomarkers

Cardiac adiposity was assessed by measuring the epicardial [16] and paracardial fat [18] on the axial view of low-dose CT images (Figure 2F). Renal adiposity was assessed at the level of the renal sinus and included combined peri-and-para-renal fat thickness (Figure 2G). Renal sinus fat was visually graded into four categories (absent-limited, low, intermediate, and high) based on previous studies [19,20] and showed good intra-observer reproducibility (Supplemental Methods). Ectopic liver fat was assessed using mean liver attenuation measured in segment VI or VII of the liver (Figure 2H), with a cut-off value of ≤48 Hounsfield Unit (HU) for hepatic steatosis [21].

#### Great vessel diameter and vascular calcification

Measurements of great vessel diameters were performed on pulmonary artery, ascending and descending aorta (Figure 2E) on the axial view of low-dose CT images. Thoracic aorta calcifications were visually graded as absent, mild, moderate, or severe based according to previously described methods [22,23]. Coronary calcifications of were visually categorized as absent-limited, moderate, or severe (Figure 2D). Great vessel diameters measurements and visual scoring of severity of calcifications of the thoracic aorta and coronary arteries on low-dose CT were compared to with measurements on ECG-gated CT and ECG-gated Agaston calcium scores and showed good agreement (Supplemental data).

#### Frailty-related biomarkers

Vertebral bone density was assessed using the average attenuation of three thoracic vertebral bodies, with cut-off values of <102 HU for low thoracic bone mineral density [24]. Para-spinal muscle density was measured at the level of T4-T7 (Figure 2I) by placing ROI while avoiding visible intramuscular of adjacent fat. Low muscle density was defined according to previously published cut-off values of <41 HU and <37 HU for men and women, respectively [25,32].

#### Renal inflammatory biomarkers

Renal inflammation marker was assessed from localized fat distribution surrounding the kidney, namely the perinephric fat stranding. Perinephric fat stranding was visually assessed and categorized into three categories (low, intermediate, and high) [26].

### Echocardiography measures and diastolic dysfunction grading

Available transthoracic echocardiographic examinations (GE Vingmed Ultrasound AS) were reviewed using Echo PAC and ViewPoint 6 (GE HealthCare). Measures of LV systolic and diastolic function, pulmonary arterial systolic pressure (PASP), and LV hypertrophy status were collected (figure 1K). LV diastolic dysfunction was graded according to the recommendation of American Society of Echocardiography (ASE) and the European Association of CardioVascular Imaging (EACVI) [27]. Classification was based on transmitral E/A ratio, septal and lateral mitral annular e’ velocity, E/e’ ratio, left atrium volume index, and tricuspid regurgitation maximum velocity [27]. Right atrial pressure was estimated from the inferior vena cava diameter and collapsibility. Echocardiographic variables required for H_2_FPEF score calculation were systematically collected. When these parameters were unavailable from routine clinical reports, measurements were retrospectively performed by a single observer using archived echocardiographic examinations according to ASE/EACVI recommendations [27] and reviewed by an experienced cardiologist (M.d.H).

### Collection of clinical data

Clinical and outcome data were extracted from electronic health records, which included demographics, cardiovascular risk factors, relevant comorbidities, and medication use. Laboratory data included markers of cardiac, and renal function, and if available of metabolic function. Estimated glomerular filtration rate (eGFR) was calculated from serum creatinine according to the CKD-EPI 2021 equation. Moreover, H2FPEF score, CKM stage, and longitudinal outcome was determined, as described below. A detailed overview of all collected variables is provided in Supplemental Table S1.

#### H_2_FPEF score

The probability of HFpEF was assessed using the H_2_FPEF score [28, 29], which incorporates age, body mass index (BMI), use of antihypertensive medication, history of atrial fibrillation, and echocardiographic measures of filling pressure (E/e’) and PASP [28]. H2FPEF score was selected because it can be readily derived from routinely available clinical and echocardiographic data [30], whereas application of the HFA-PEFF algorithm was limited by the lack of systematically available B-type natriuretic peptide.

#### Cardio-Kidney-Metabolic syndrome stage

CKM stages were determined according to the 2026 AHA framework using available clinical, laboratory, and imaging-derived data [31]. Staging incorporated adiposity-related risk factors, metabolic abnormalities, CKD, subclinical and established cardiovascular diseases. Briefly, stage 0 indicates the absence of CKM risk factors, stage 1 the presence of excess of dysfunctional adiposity, stage 2 the presence of metabolic risk factors and/or chronic kidney disease, stage 3 subclinical cardiovascular (CV) disease or elevated predicted CV risk, and stage 4 established CV disease. Stage assignment was based on all available information from medical history, laboratory testing, echocardiography, and imaging examinations.

### Longitudinal Outcome

Information on mortality and MACE is prospectively collected from clinical records, with MACE being defined as a 5-point composite outcome compromising cardiovascular death, myocardial infarction, stroke, heart failure hospitalization, and coronary revascularisation. Coronary revascularization performed within 6 months after cardiac ²Rb-PET/CT was excluded from the MACE endpoint if the procedure was initiated as a direct consequence of the PET/CT findings. Moreover, new occurrence of atrial fibrillation was prospectively collected.

### Adjustment for heigh-related selection bias

In the APOLLO registry, individuals of shorter stature were overrepresented because inclusion required sufficient kidney coverage within the scan range of the ²Rb-PET/CT examination to derive both myocardial and renal perfusion measurements from the same scan. When extrapolating findings to a population with an unselected height distribution, this overrepresentation may introduce selection bias due to the skewed height distribution in the APOLLO registry. Weighting the study population to reflect the height distribution of the unselected population may mitigate this bias [12,13]. Therefore, a random sample of 504 unselected patients who underwent ²Rb-PET/CT imaging at Alrijne Hospital during the same period was used as a reference population to calculate sampling weights for the APOLLO registry using inverse probability of sampling weighting [Figure 1]. This approach resulted in higher sampling weights for taller participants, who were underrepresented in the registry. Applying these weights allows estimates to better reflect the population undergoing ²Rb-PET/CT imaging without height-related oversampling. Supplementary Figure 2 demonstrates that applying the sampling weights resulted in a height distribution comparable to that of the unselected reference population.

### Limitations

Several limitations should be acknowledged. First, the retrospective nature of this registry resulted in incomplete availability of clinical and imaging data. This is particularly relevant to CKM stage 2 and 3, in which incomplete ascertainment of subclinical cardiovascular disease may result in under classification. Second, the registry comprises of patients referred for clinically indicated cardiac perfusion imaging and therefore does not represent the general population, introducing potential referral and selection bias. Third, inclusion required sufficient kidney coverage within the ²Rb-PET/CT scan range to derive both myocardial and renal perfusion measurements from the same scan, resulting in an overrepresentation of shorter individuals. To account for this, inverse probability of sampling weights were calculated using a random reference sample of unselected patients who underwent ²Rb-PET/CT at the same hospital during the same period. Finally, CT-derived CKM imaging biomarkers represent emerging biomarkers for which standardization, clinical interpretation, and prognostic validation continue to evolve.

In conclusion, the APOLLO registry aims to extend data collection of cardiac 82Rb-PET/CT beyond myocardial perfusion imaging by integrating cardio-renal perfusion measurements with detailed cardiometabolic, clinical, and outcome phenotypes. This real-world registry provides a unique platform for future mechanistic and outcome-based research across the CKM disease spectrum.

## Data Availability

The data underlying this article will be shared on reasonable request to the corresponding author.

## Acknowledgement

The authors acknowledge the technical assistance and supervision given by senior nuclear medicine technologist, Mrs. Petra Dibbets-Schneider, at Leiden University Medical Center, Radiology and Nuclear Medicine Department.

## Supplemental data

**Table S1.** All data collected from medical history and imaging-derived parameters collected for APOLLO study. (maybe supplemental)

| No | Categories | Subgroup | Descriptions |
| --- | --- | --- | --- |
| 1 | Demographics |  | Age, sex |
| 2 | Imaging modalities |  | <sup>82</sup>Rb-PET/CT, cardiac-gated CT, TTE |
| 3 | Electrocardiogram (ECG) |  | 12-lead ECG during rest and stress |
| 4 | Blood pressure |  | Systolic, diastolic, mean arterial pressure |
| 5 | H2FpEF score components |  | Age, BMI, E/e', Pulmonary arterial systolic pressure, Atrial fibrillation, >2 HT medications |
| 6 | Medical history | Cardiometabolic risk factors | Hypertension, DM, CKD, smoking status, BMI |
|  |  | Atherosclerotic CVD | Myocardial Ischemia, Coronary Artery Disease, Stroke/ Transient Ischemic Attack, Peripheral Artery Disease |
|  |  | Cardiac rhythm and conduction disorder | Atrial Fibrillation, Arrhythmias, Conduction disorder |
|  |  | Pulmonary comorbidity | COPD, Asthma, Pulmonary hypertension, Obstructive sleep apnoea syndrome |
|  |  | Cardiac vascular interventions | CABG, PCI, pacemakers |
| 7 | Medications | Anti-ischemic | Nitrates, β-blockers, Calcium channel blockers, |
|  |  | Anti-thrombotic | Anti-platelets, Anti-coagulation |
|  |  | Lipid-lowering | Statins, Ezetimibe |
|  |  | Anti-hyperglycaemic | Insulin, GLP-1 receptor agonist, SGLT2i, metformin |
|  |  | RAAS modulation | ACEi, ARBs, Aldosterone antagonists |
|  |  | Heart Failure | Diuretics |
| 8 | PET-derived hemodynamic biomarkers | Cardiac | Heart rate (stress, rest), MBF (stress, rest), MFR, iMFR LVEDV, LVESV, LVEF, TID |
|  |  | Renal | Renal Blood Flow, Effective Renal Plasma Flow, K1 |
| 9 | Low dose CT-derived biomarkers | Body composition | Epicardial and Paracardial fat thickness, Liver density, renal sinus fat (visual grade), peri-pararenal fat width, perinephric fat stranding (visual grade), thoracic vertebral bone density, paraspinal muscle density |
|  |  | Vascular | Diameter of pulmonary artery, ascending aorta, proximal descending aorta, thoracic aorta calcification (visual grade), coronary calcification (visual grade) |
| 10 | Cardiac gated non-contrast CT | Coronary calcium scoring | Agatston score (total, LAD, LCX, RAD) |
| 11 | Trans Thoracic Echocardiography (TTE) | Diastolic function | Average E/e', Mitral valve e', Tricuspid Regurgitation velocity, LV ejection fraction, Left Atrial Volume index, E/A, E & A wave, Global longitudinal strain |
|  |  | Cardiac Morphology | Relative Wall Thickness, Left Ventricle Mass index, Intra Ventricular Septum thickness |
|  |  | Right-heart function | Inferior Vena Cava diameter & collapsibility, Pulmonary Arterial Systolic Pressure, S' |
| 12 | Laboratory values | Kidney function | Serum Creatinine, eGFR (CKD-EPI) |
|  |  | Glycaemic markers | Glucose, HbA1C |
|  |  | Cardiac biomarkers | NT-proBNP |
|  |  | Hematological parameters | Hemoglobin, Hematocrit, |
|  |  | Inflammatory marker | C-reactive protein |

*Abbreviations: BMI: Body Mass Index, HT: Hypertension, DM: Diabetes Mellitus, TTE: TransThoracic Echocardiography, CKD: Chronic Kidney disease, COPD: Chronic Obstructive Pulmonary Disease, CABG: Coronary Artery Bypass Graft, PCI: Percutaneous Coronary Intervention, RAAS: Renin-Angiotensin-Aldosterone System, ACE: Angiotensin Converting Enzyme, ARB: Angiotensin Receptor Blockers, MBF: Myocardial Blood Flow, MFR: Myocardial Flow Reserve, iMFR: integrated myocardial flow reserve, LVEDV: Left Ventricle End Diastolic Volume, LVESV: Left Ventricular End Systolic Volume, LVEF: Left Ventricle Ejection Fraction, TID: Transient Ischemic Dilation, LAD: Left Anterior Descending, LCx: Left Circumflex, RCA: Right Coronary Artery, eGFR: estimated Glomerulus Filtration Rate, LDL-C: Low-Density Lipoprotein Cholesterol, HDL-C: High-Density lipoprotein Cholesterol, NTproBNP: N-Terminal pro B-type Natriuretic Peptides*

### Imaging acquisition

Rubidium-82 PET/CT scans were acquired using a Discovery MI 5-ring PET/CT (GE HealthCare, Chicago, Illinois, USA) (figure 2A). A low dose CT (20 mAs, 120kV) was acquire for attenuation correction purposes, with 512 x 512 matrix (1.4 x 1.4 x 3.75 mm^3^) and field of view of 70 cm. A 10-leads ECG ST80i stress testing system (Phillips HealthCare, Best, The Netherlands) was utilized for cardiac gating. In case additional calcium scoring was performed, a ECG-gated CT scan was acquired (100 mA, 120kV, 0.35 s rotation, axial thickness of 2.5 mm, and 16 images per rotation). PET acquisitions were performed for 6 minutes at rest and during pharmacological stress induced by either adenosine (140µg/kg/min) or a single bolus of regadenoson (400µg). Image acquisitions started approximately 8 sec after automated intravenous infusion of 750 MBq ^82^RbCl (^82^SR/^82^Rb generator, Rubigen B.V, the Netherlands). PET data were acquired in list mode and reconstructed into dynamic frames using time-of-flight ordered subset expectation maximization algorithm (VPFX TOF-OSEM:; 2 subsets, 17 iterations) on a 128 by 128 matrix (2.8 x 2.8 x 2.8 mm^3^) with a 5.0 mm full-width-at-half-maximum Hanning filter. A total of 26-time frames were reconstructed per PET scan, that is, 10 × 5, 6 × 10, 5 × 15, and finally 5 × 20 s. Images were normalized and corrected for radioactive decay, attenuation, and scatter [1].

**Figure S1.**
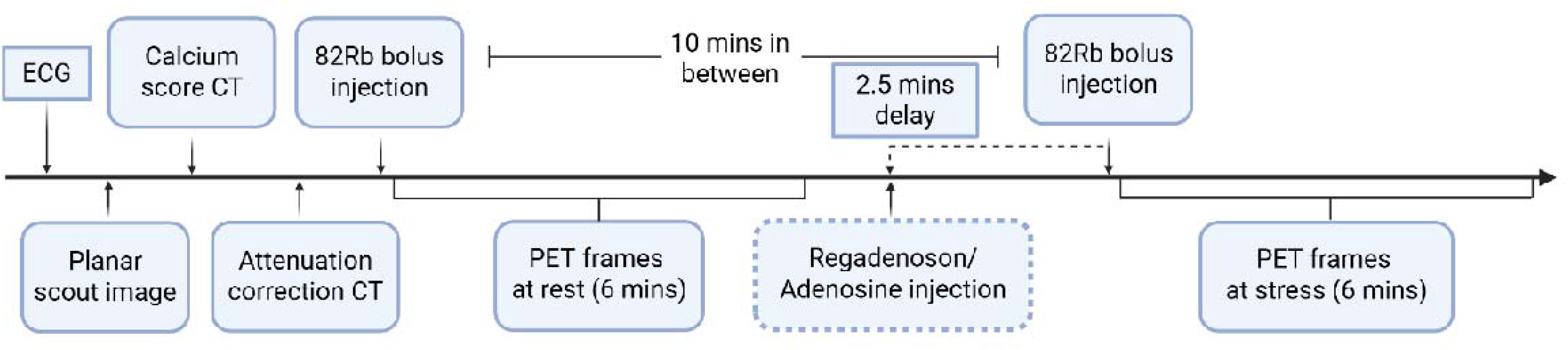
Overview of Imaging acquisition of APOLLO registry. ECG: Electrocardiogram, CT: Computed Tomography, PET: Positron emission tomography, 82Rb: Rubidium - 82

### Collection of PET-derived hemodynamic measures

#### Cardiac perfusion

Patients’ dynamic PET/CT scans at rest and during stress were analysed using Corridor4DM software (v2024.0.0.174 ngen, INVIA Medical Imaging Solution, Ann Arbor, MI, USA) (Figure S2A). LV was automatically segmented and manually corrected as necessary under the supervision of an experienced nuclear medicine technologist (P.D-S). Myocardial blood flow was estimated by fitting a one-tissue-compartment kinetic model to myocardial time activity curve, with myocardial flow reserve defined as ratio of myocardial flow at rest and during stress [2]. Reversibility map of myocardial perfusion was averaged over the whole LV and visualized using 17 segments bulls eye plot [2,3] of LV to reflect perfusion defect and severity. In addition, myocardial perfusion imaging and myocardial flow reserve (MFR) were also assessed to create integrated myocardial flow reserve (iMFR) map of the LAD, LCx, RCA, and global perfusion. The categorisations of iMFR are defined as: normal myocardium with MFR >2.0 and no relative perfusion defect, diffusely impaired with MFR <2.0 and no relative perfusion defect, focally preserved myocardium with MFR>2.0 and relative perfusion defect, and focally impaired myocardium with MFR <2.0 and relative perfusion defect. The threshold of 2.0 for MFR was derived from previous studies, which has demonstrated the ability of iMFR to predict cardiac-related morbidity and mortality [2,3]. In this framework, diffusely impaired flow (yellow) was interpreted as coronary microvascular dysfunction, whereas focally impaired flow (red) was interpreted to reflect a predominantly epicardial (obstructive) pattern. This iMFR classification was used to distinguish microvascular from epicardial disease, allowing patients with obstructive coronary artery disease to be retained rather than excluded. MFR will be corrected to normalize effect of hypertension and tachycardia to MBF at rest by dividing MBF at rest with rate pressure product (corrected MBF at rest= MBF at rest / [heart rate x systolic blood pressure]), thus, MFR corrected = MBF during stress/ corrected MBF at rest, as previously described. Bland-Altman analysis was utilized to measure the mean bias and 95% limits of agreement between two independent observers for the measurements of MFR (0.0205, -0.3807 to 0.4217), MBF during stress (0.0720, -0.3213 to 0.0790) and MBF at rest (-0.0035, -0.0860 to 0.0790) (Figure S2)

**Figure S2.**
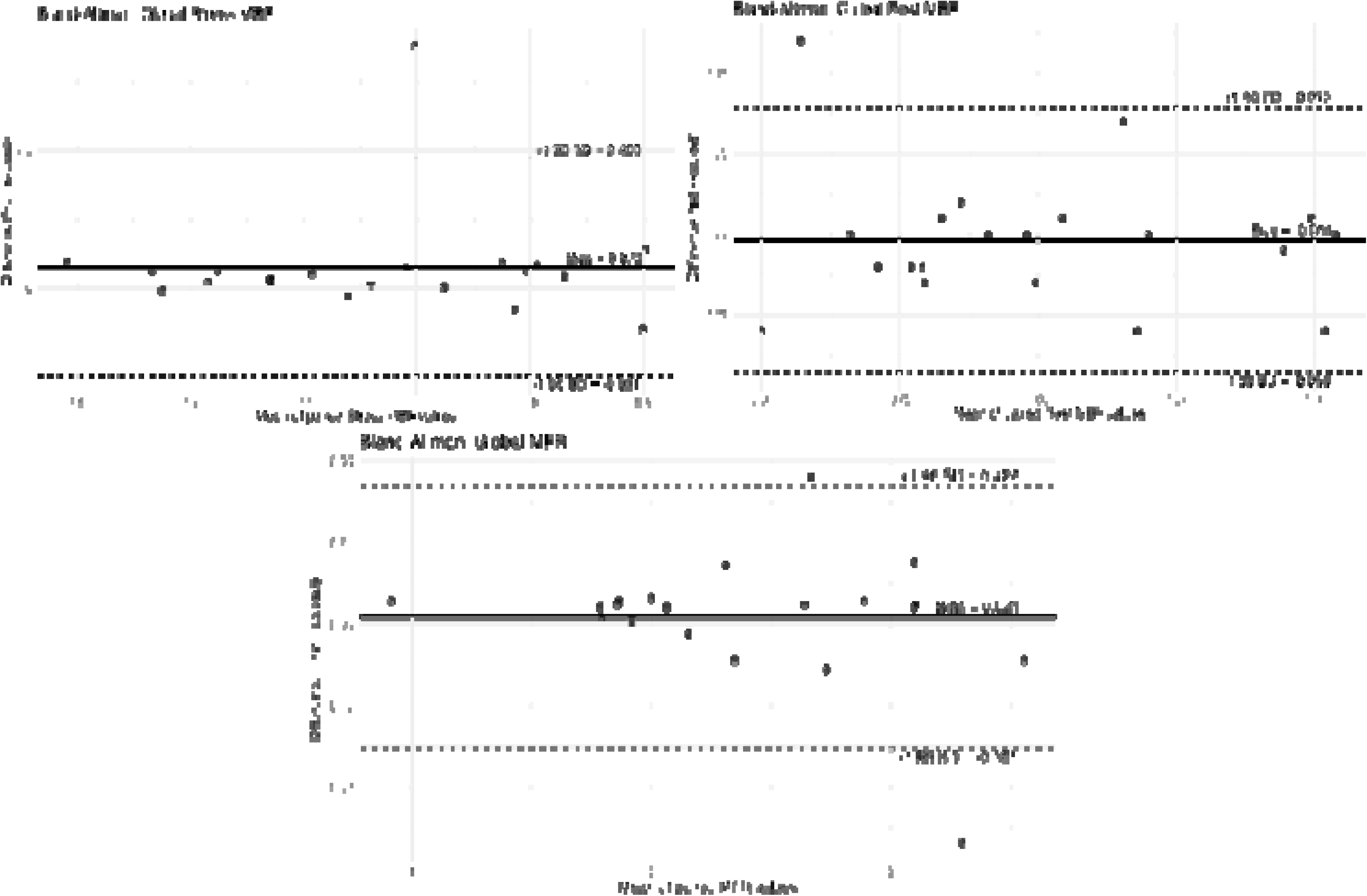
Bland-Altman analysis for cardiac perfusion measurements between two observers for Global Myocardial blood flow at stress, rest, and myocardial flow reserve. Mean bias and 95% limits of agreement are indicated in each plot. A) Inter-observer variability between two independent observers for MBF measurement at rest. (B) Inter-observer variability between two independent observers for MBF measurement at rest. (C) Inter-observer variability with two independent observers for MFR. MBF: myocardial blood flow, MFR: myocardial flow reserve

#### Renal perfusion

An existing one-tissue compartment model (Figure S3) was used for quantification of renal blood flow (RBF) utilizing kinetic modelling feature of PMOD software package (version 4.2; PMOD Technologies Ltd, Zurich, Switzerland).

**Figure s3.**
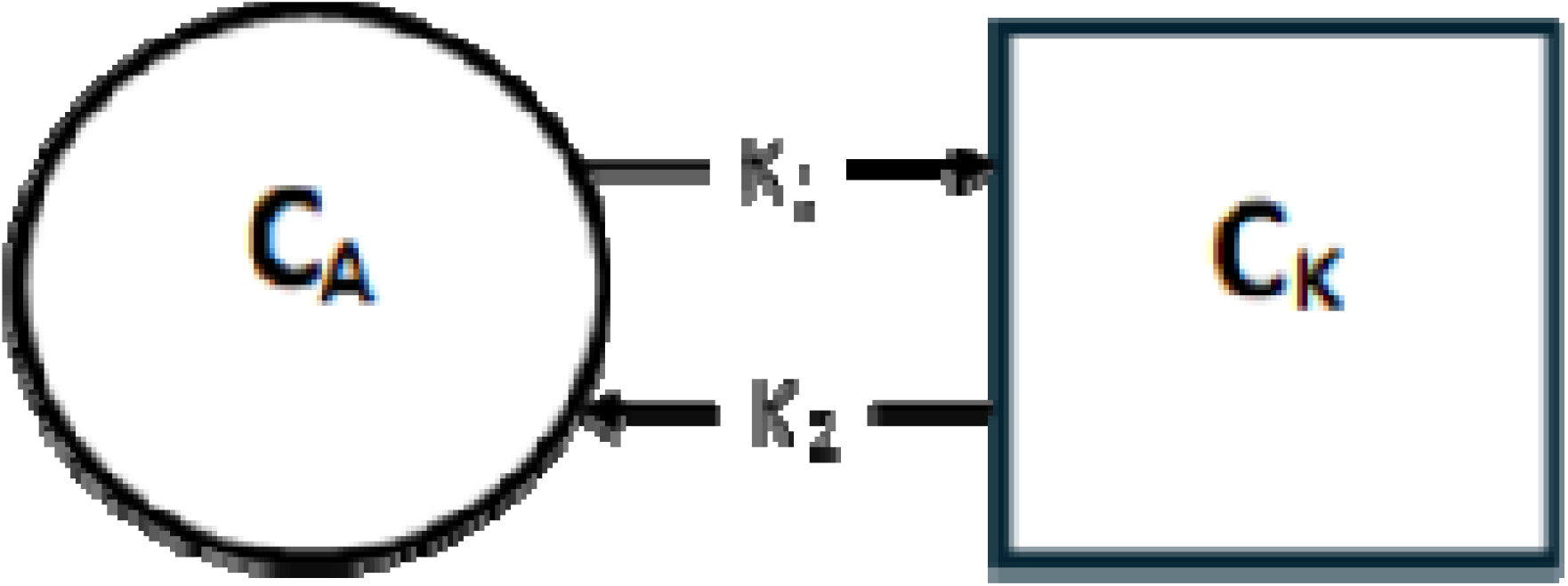
One-tissue compartment model used for RBF quantification. C_A_: vascular compartment (kBq/mL) as input function, C_K_: Kidney compartment (kBq/mL), K_1_: uptake rate of ^82^Rb from vascular to kidney compartment, K_2_: fraction of ^82^Rb from kidney back to vascular compartment.

Dynamic ^82^Rb-PET/CT images at rest were used to delineate a volume-of-interest (VOI) on the cortex of superior kidney slice (C_K_) and in the abdominal aorta (C_A_). To limit the effect of spill-over and partial volume, VOI was placed >2 voxels away from the organ’s boundary walls. VOI did not include the renal pelvis and was not placed in three most superior or inferior slices of the kidney, due to reduced spatial resolutions [1]. PMOD was used to create an operational model (1), with blood volume fraction (V_B_) considered to have a constant value of 0.10 [1,4]. The rate of constant K_1_ and K_2_ were estimated by PMOD. A correction for delay and dispersion of the blood signal was done to increase accuracy of arterial concentration measurement, and was integrated into one-tissue compartment model 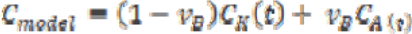 (1) After the VOI placement in renal cortex and abdominal aorta, time activity curve was generated to reflect the mean activity concentrations of ^82^Rb (kBq/ml) over time (figure 2C-right). K_1_ value was converted to RBF using equation 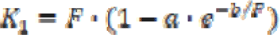 (2), with assumptions: constant extraction fraction (*e*) of 0.9, (*a*) of 0.94, (*b*) of 1.86, and conversion of flow values to mL/min/g with kidney density of 1.04g/ml [1,4,5]. Bland-Altman analysis was utilized to measure the absolute agreement between two independent observers for the measurements of RBF at rest and intra-observer repeatability of 30 repeated RBF measurements (Figure S4).

**Figure s4.**
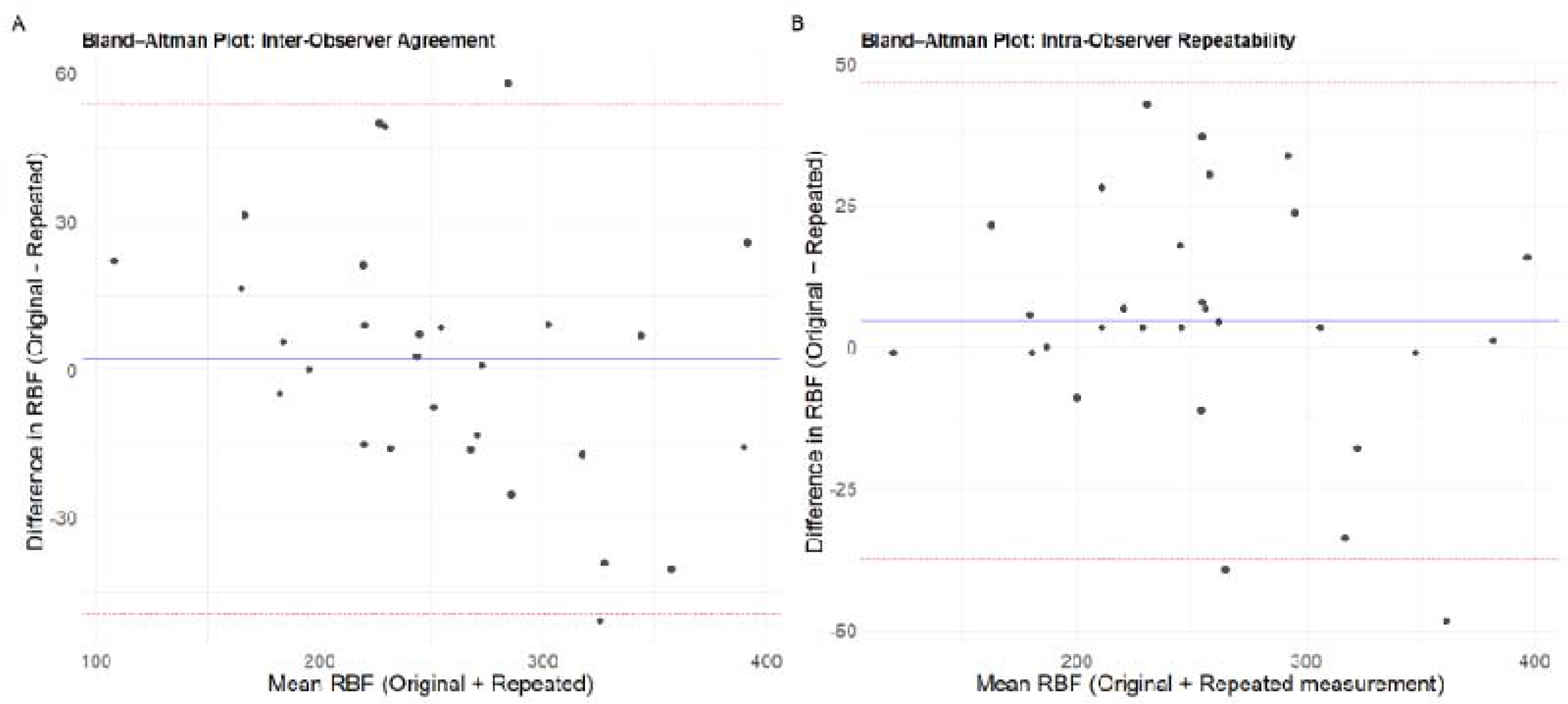
Bland-Altman agreement of Renal Blood Flow measurement at rest for inter-observer agreement between two independent analysts and intra-observer repeatability for 30 measurements, measured using PMOD. Mean bias and 95% limits of agreement are indicated in each plot. A) Inter-observer variability with two independent observers, blinded to the original measurements and re-analysed the same scans. (B) Intra-observer variability, primary analyst repeated 30 RBF measurements at a separate time point and blinded to the original results.

### Great Vessel Diameter

Measurements of great vessel diameter were performed using the low-dose CT. Due to the known effect of change in diameter during the cardiac cycle, these measurements introduced potential error for pulmonary artery, ascending, and descending aorta [17]. Therefore, a reproducibility analysis was performed to measure the difference in diameter measurements between low-dose CT and cardiac-gated CT in 30 randomly selected individuals. Bland-Altman analysis was utilized to measure the mean bias and 95% limits of agreement between low-dose and cardiac-gated, which demonstrated comparable agreement for diameter measurement of ascending aorta (-0.80, -3.21 to 1.23) descending aorta (-0.13, -1.94 to 1.68), and pulmonary artery (-0.60, -4.19 to 2.99) (Figure S5).

**Figure s5.**
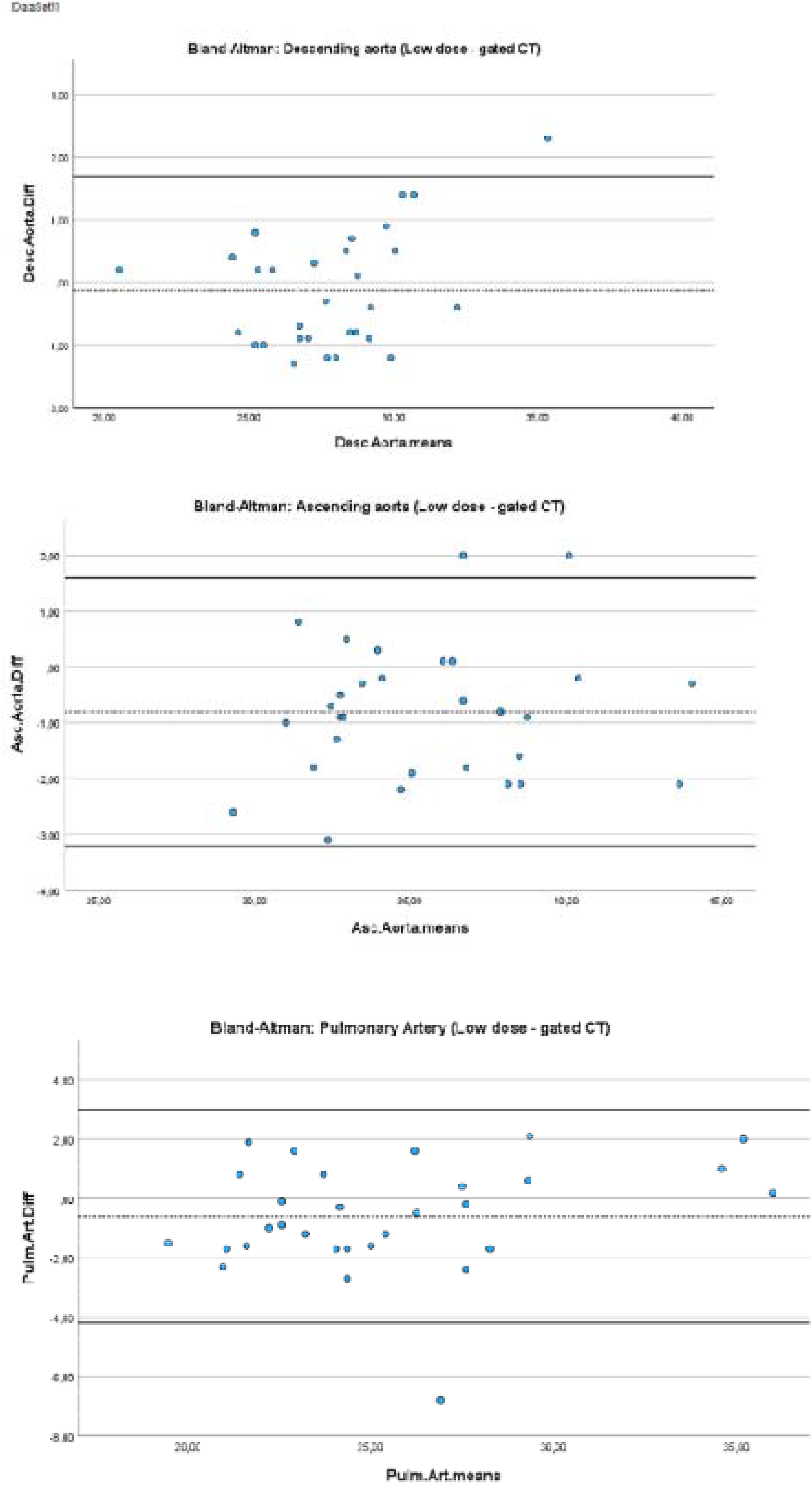
Bland-Altman agreement of diameter measurement for ascending aorta, descending aorta, and pulmonary artery between low-dose CT and cardiac-gated CT. Mean bias and 95% limits of agreement are indicated in each plot. (A) Inter-modality variability of ascending aorta diameter measurement between low-dose and cardiac-gated CT. (B) Inter-modality variability of descending aorta diameter measurement between low-dose and cardiac-gated CT. (C) Inter-modality variability of pulmonary artery diameter measurement between low-dose and cardiac-gated CT.

**Table s2.**
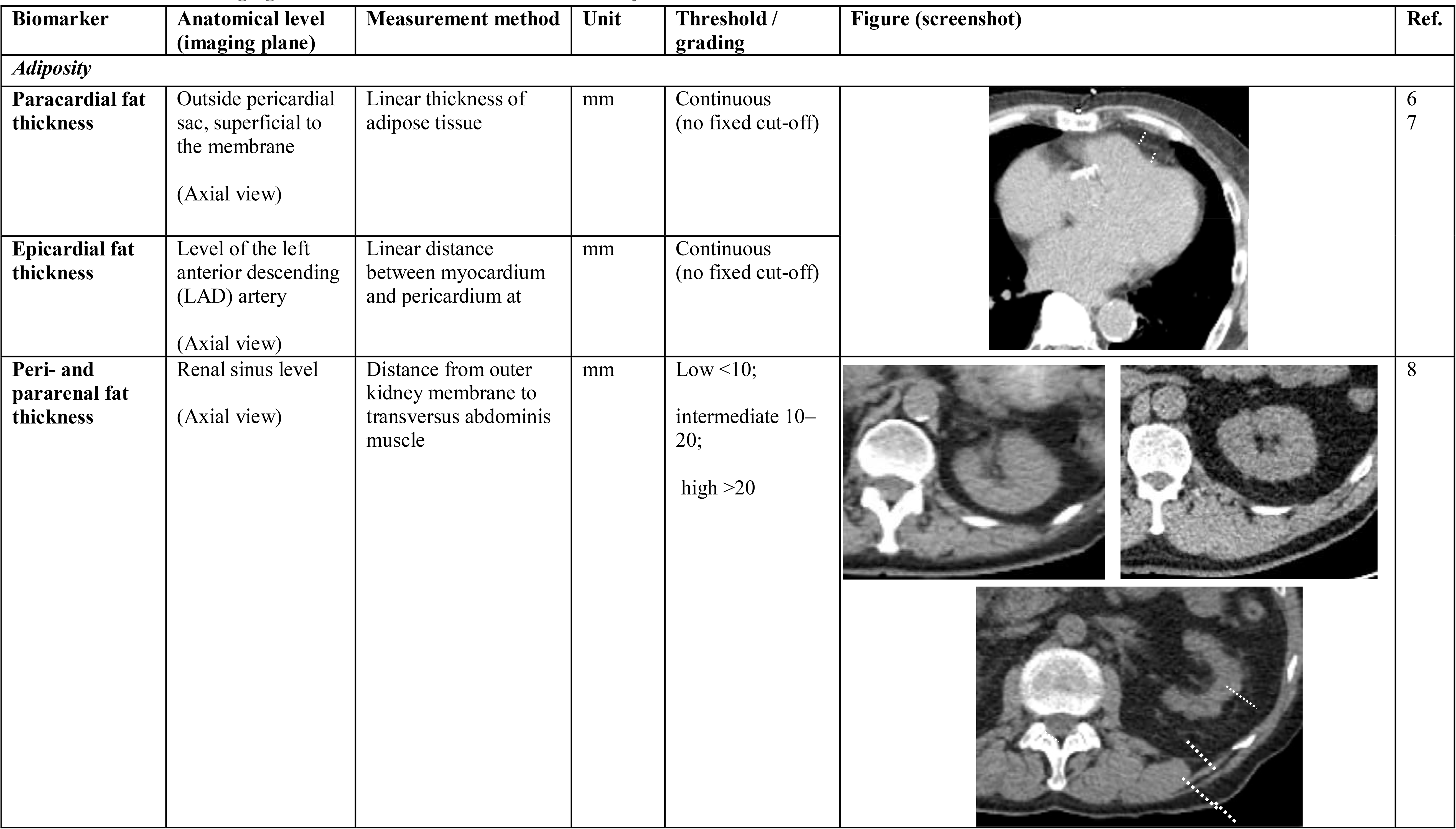
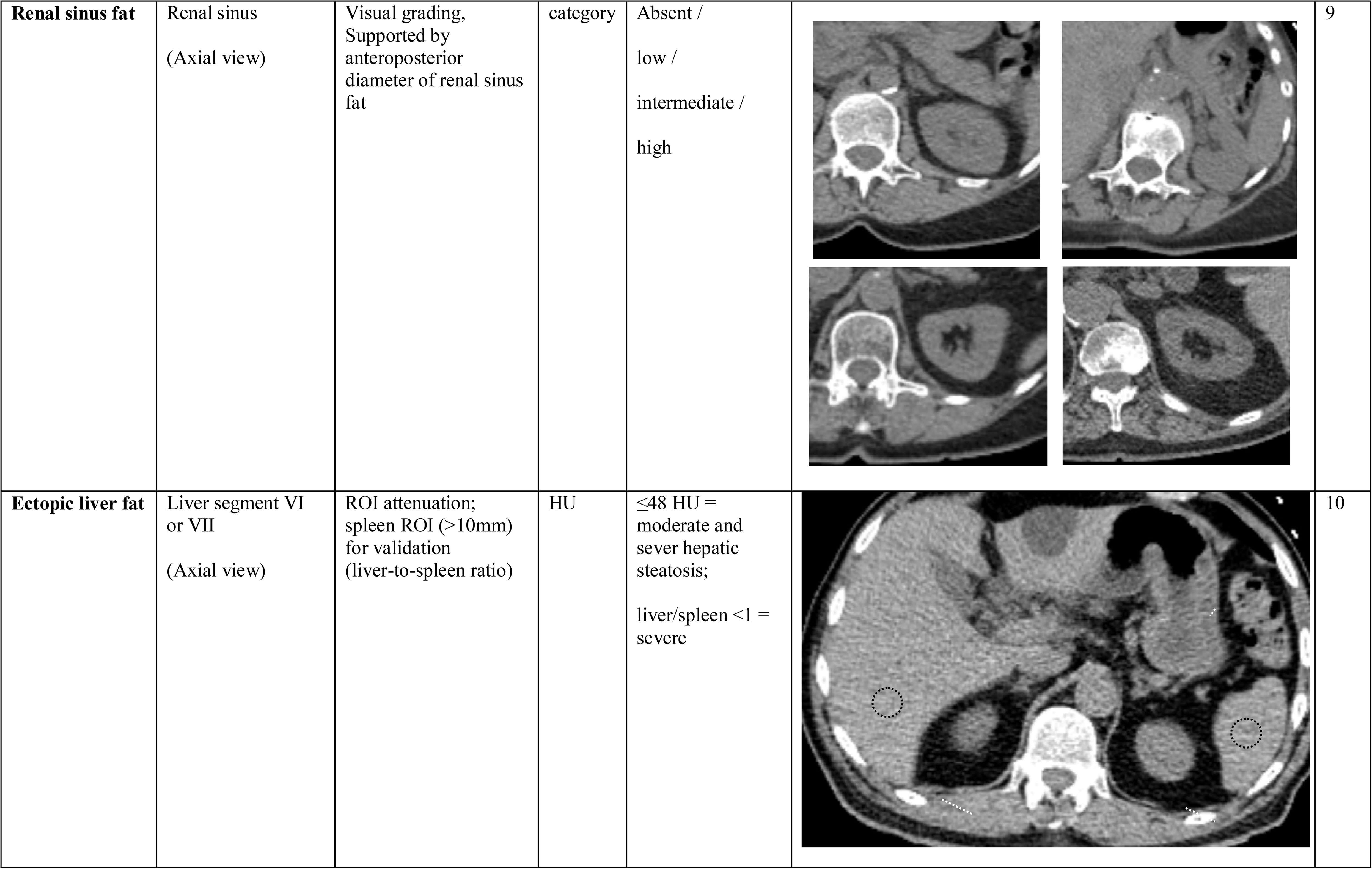
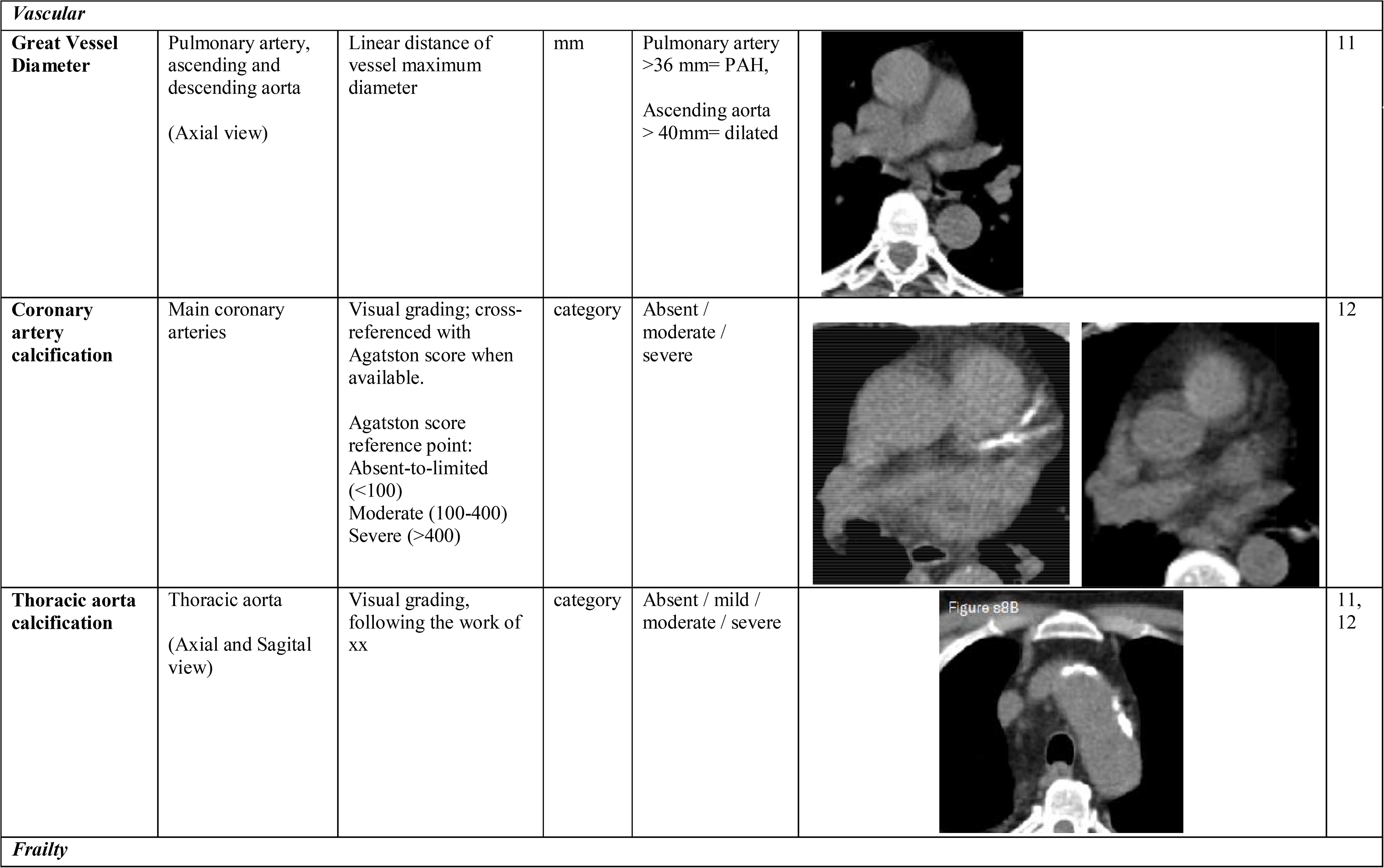
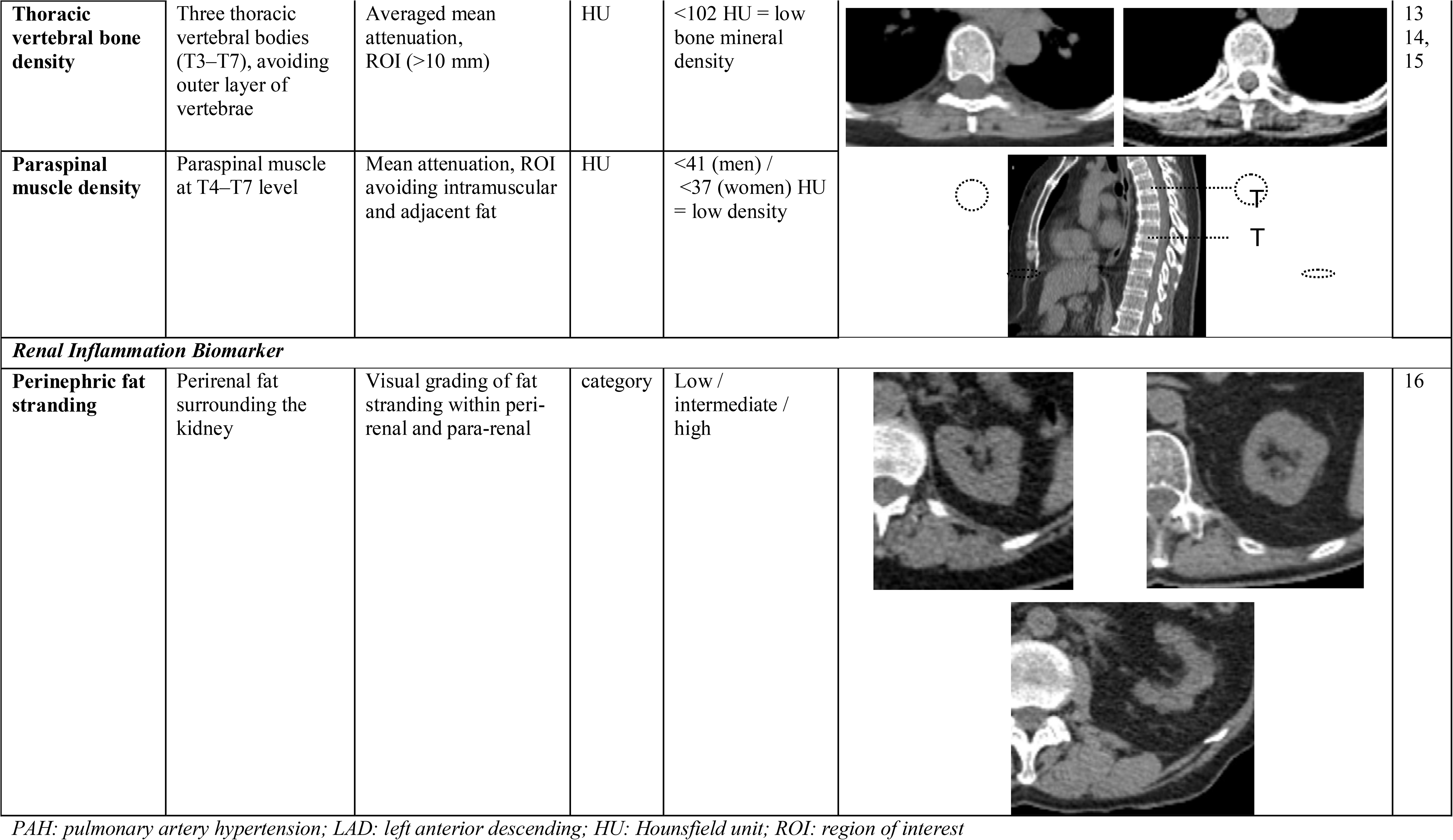
CT-derived imaging biomarkers collected for APOLLO study.

### Inverse Probability of Selection Weighting analysis

The requirements for a sufficient renal slice within the PET/CT scan led to oversampling of shorter individuals. In addition, differences in age and sex may have also influenced the likelihood of being included. Inverse probability of selection weighting analysis (IPSW) was performed to assess potential impact of differential selection into the final imaging cohort. For the weighted analysis, the reference population was comprised of individuals eligible for the registry but were not included in the final cohort due to inadequate renal coverage (Figure 1). To account for temporal effects, 14 individuals were randomly selected for each month between 2021 and 2023, totalling of 504 individuals in the reference populations.

The probability of cohort inclusion was estimated using a multivariable logistic regression model incorporating prespecified characteristics considered potentially relevant to selection, including age, sex, and height. The distributions of these parameters before and after weighting were assessed to evaluate whether IPSW reduced differences related to the selection mechanism (Figure S6). The predicted probability of inclusion represented each individual’s propensity for selection into the final cohort (supplemental excel). Consistency of effect estimates and their confidence intervals between weighted and unweighted analyses was used to assess the robustness of the findings to potential selection related to the measured characteristics. IPSW does not account for selection factors that were not measured or included in the weighting model; therefore, residual selection bias cannot be excluded.

**Figure S6.**
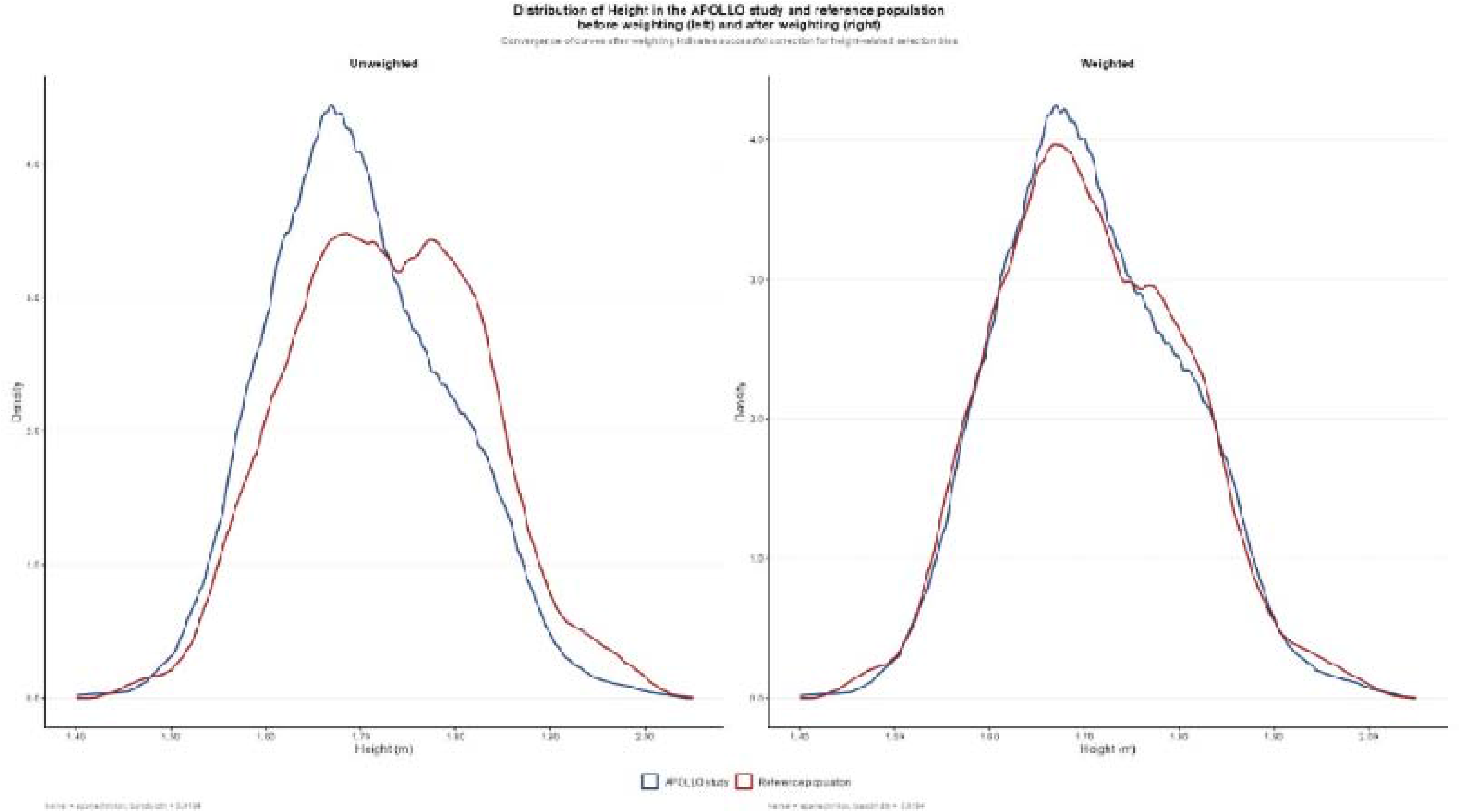
Distribution of height in the APOLLO study and reference population before and after IPSW analysis/weighting. Kernel=epanechikov, bandwidth= 0.0194.

